# Diagnosing pulmonary tuberculosis using sequence-specific purification of urine cell-free DNA

**DOI:** 10.1101/2021.01.19.21249296

**Authors:** Amy Oreskovic, Nuttada Panpradist, Diana Marangu, M. William Ngwane, Zanele P. Magcaba, Sindiswa Ngcobo, Zinhle Ngcobo, David J. Horne, Douglas P.K. Wilson, Adrienne E. Shapiro, Paul K. Drain, Barry R. Lutz

## Abstract

Transrenal urine cell-free DNA (cfDNA) is a promising tuberculosis (TB) biomarker, but is challenging to detect because of the short length (<100 bp) and low concentration of TB-specific fragments. We aimed to improve the diagnostic sensitivity of TB urine cfDNA by increasing recovery of short fragments during sample preparation. We developed a highly sensitive sequence-specific purification method that uses hybridization probes immobilized on magnetic beads to capture short TB cfDNA (50 bp) with 91.8% average efficiency. Combined with short-target PCR, the assay limit of detection was ≤5 copies of cfDNA in 10 mL urine. In a clinical cohort study in South Africa, our urine cfDNA assay had 83.7% sensitivity (95% CI: 71.0– 91.5%) and 100% specificity (95% CI: 86.2–100%) for diagnosis of active pulmonary TB when using sputum Xpert MTB/RIF as the reference standard. The detected cfDNA concentration was 0.14–2804 copies/mL (median 14.6 copies/mL) and was inversely correlated with CD4 count and days to culture positivity. Sensitivity was non-significantly higher in HIV-positive (88.2%) compared to HIV-negative patients (73.3%), and was not dependent on CD4 count. Sensitivity remained high in sputum smear-negative (76.0%) and urine LAM-negative (76.5%) patients. With improved sample preparation, urine cfDNA is a viable biomarker for TB diagnosis. Our assay has the highest reported accuracy of any TB urine cfDNA test to date and has the potential to enable rapid non-sputum-based TB diagnosis across key underserved patient populations.

## INTRODUCTION

Tuberculosis (TB) is the leading cause of global mortality due to infectious disease, with an estimated 10 million cases and 1.4 million deaths in 2019 (1). An estimated 30% of TB cases remain undiagnosed or unreported, in part due to limitations in rapid diagnostics (1). Current TB tests rely on sputum samples, which are difficult to collect from people living with HIV, severely ill patients, and children, and may not detect extrapulmonary TB (EPTB). Rapid sputum-based tests (*e*.*g*., smear microscopy, Xpert MTB/RIF) also have lower sensitivity in these same underserved patient populations, who more often have paucibacillary TB (2–4). A WHO consensus meeting identified a rapid, non-sputum-based test as one of the highest priority target products for TB diagnostics (5).

Urine is an attractive alternate sample for TB diagnosis because it is easy to collect and poses minimal transmission risk. In patients with active TB disease, TB-specific cell-free DNA (cfDNA) fragments are released into the blood, a fraction of which are filtered through the kidneys and excreted in the urine as transrenal cfDNA (6–8). TB-specific cfDNA has been detected in urine from both HIV-negative and HIV-positive patients with pulmonary TB, but diagnostic sensitivities have been inconsistent (0 – 79%) (8–13). High variability in methodology and subsequent performance across studies have limited the understanding of TB urine cfDNA and hindered its clinical implementation (14, 15).

Urine cfDNA is a challenging target due to the short length and low concentration of TB-specific fragments. While plasma cfDNA has a peak fragment length of 167 bp, urine cfDNA is more fragmented (7, 16, 17). Recently, new sequencing library preparation methods revealed that very short, formerly-undetectable fragments compose a larger fraction of cfDNA than previously realized (18–21). The majority of urine cfDNA fragments are <100 bp, with peak fragment length ranging from 30 – 110 bp (16, 17, 21, 22). Although the fragment length of TB cfDNA has not yet been characterized, bacterial cfDNA is expected to be especially fragmented (peak <60 bp) because it is less protected by DNA-associated proteins (21, 23).

The low to moderate sensitivities reported in previous TB urine cfDNA studies are likely due in part to sample preparation and/or amplification methods that are sub-optimal for short urine cfDNA (6, 14). We hypothesized that we could increase the sensitivity of cfDNA-based TB diagnosis by improving recovery of short cfDNA during the DNA extraction step. We developed a highly sensitive sequence-specific purification method that uses DNA probes immobilized on magnetic beads to extract TB-specific cfDNA via hybridization. By combining sequence-specific purification with short-target PCR, we can reliably detect ≤5 copies of short (50 bp) cfDNA in 10 mL urine (24).

Here, we determined the diagnostic accuracy of our TB cfDNA assay in clinical urine specimens for the first time. Our results demonstrate the advantages of our sequence-specific purification approach, contribute to the growing evidence needed to establish urine cfDNA as a TB biomarker, and will serve as the foundation for future clinical studies in expanded populations (*e*.*g*., children, individuals with EPTB).

## MATERIALS AND METHODS

### Study design and participants

Participants were enrolled at Edendale Hospital in Pietermaritzburg, South Africa (approved by the University of KwaZulu-Natal Biomedical Research Ethics Committee, #BE475/18). Adults (≥18 years old) meeting the case definition for active pulmonary TB were recruited. As a control group, adults with excluded TB diagnosis and no suspicion for active TB disease were enrolled. A separate healthy control group was enrolled at the University of Washington, Seattle, USA (approved by the University of Washington Institutional Review Board, #48840). Participants were adults (≥18 years old) with low risk of TB exposure, as defined by birth in a country with low TB risk and no history of diagnosis of latent TB, treatment for active or latent TB, or living with an individual with active TB. All participants provided written informed consent. Samples were de-identified prior to testing at the University of Washington where this study was conducted.

### Case definitions

TB-positive participants (South Africa) were defined as those with a positive sputum Xpert MTB/RIF (Cepheid, Sunnyvale, CA, USA) result within 5 days of enrollment and the presence of one or more TB symptom (fever, night sweats, cough, and weight loss). Patients with >72 hours of anti-TB treatment were excluded. TB-negative participants (South Africa) had a primary diagnosis other than TB and no clinical suspicion for TB. Healthy controls (USA) were recruited on a volunteer basis and were not seeking medical care.

### Clinical data, sputum testing, and urine lipoarabinomannan (LAM) testing

Clinical data were collected for all participants enrolled in South Africa. These included date of birth, gender, weight, height, self-reported TB history, presence of TB symptoms (fever, night sweats, cough, weight loss), TB treatment duration, HIV test result, and CD4^+^ cell count (for participants living with HIV only). Expectorated sputum was submitted to the South African National Health Laboratory System (NHLS) for confirmatory solid and liquid mycobacterial culture and AFB smear microscopy. Mycobacterial culture was performed at the NHLS Provincial TB Reference Laboratory using Middlebrook 7H11 solid agar medium and the liquid BACTEC mycobacterial growth indicator tube (MGIT) 960 system (BD, Franklin Lakes, NJ, USA). Cultures were incubated for up to 42 days. Culture plates were read at 3 and 6 weeks, and *M. tuberculosis* was identified from solid or liquid cultures using niacin and nitrate testing. Participants were considered culture-positive with growth from either the solid or liquid culture. Smear microscopy was performed in the NHLS Laboratory at Edendale Hospital on decontaminated samples using both Ziehl-Neelson and Auramine stains and considered positive if either stain revealed AFB. Urine was tested using Alere Determine TB LAM Ag (Abbott Laboratories, Chicago, USA). Collection of clinical data, sputum testing, and urine LAM testing were not done for healthy controls enrolled in the USA.

### Urine collection and storage for cfDNA analysis

Each participant was asked to provide a urine sample upon enrollment. As soon as possible after collection, urine was mixed with EDTA and Tris-HCl pH 7.5 to final concentrations of 25 mM and 10 mM, respectively. Urine was stored in DNA LoBind tubes (Eppendorf, Hamburg, Germany) at -20°C at the collection site until shipping. Samples were shipped on dry ice to the University of Washington, where they were stored at -80°C until analysis. Immediately before analysis, urine was thawed at 37°C and centrifuged at 8,000g for 5 minutes to pellet cell debris. The cell-free urine supernatant was transferred to new 15 mL DNA LoBind tubes (Eppendorf) and characterized using Fisherbrand 10-SG Urine Reagent Strips (Thermo Fisher Scientific, Waltham, MA, USA) to measure the levels of glucose, bilirubin, ketone, specific gravity, blood, pH, protein, urobilinogen, nitrite, and leukocytes.

### Purification of TB cfDNA from urine sequence-specific hybridization capture

Urine (10 mL) was analyzed in triplicate for each participant, with individual replicates processed on separate days. TB-specific urine cfDNA was extracted using our in-house sequence-specific hybridization capture method, as described previously (24). We have published a detailed, user-ready protocol at http://dx.doi.org/10.17504/protocols.io.bep4jdqw.

#### Immobilization of capture probes on magnetic beads

Dynabeads MyOne Streptavidin C1 (Thermo Fisher) were washed three times with an equal volume of high salt wash buffer (1M NaCl, 10 mM Tris-HCl pH 8.0, 0.05% (v/v) Tween-20) and resuspended in an equal volume of high salt wash buffer. Dual biotinylated capture probes BP1 and BP2 (Table 1), targeting opposite strands of the double-stranded IS*6110* target region, were pre-mixed in low-EDTA TE buffer to a concentration of 50 µM each. 25 pmol of each probe (0.5 µL of probe mix) per 50 µL bead equivalent was added to the beads. The beads were immediately vortexed and rotated for 15 minutes at room temperature to immobilize capture probes on the beads. The beads were washed three times with an equal volume of high salt wash buffer and resuspended in an equal volume of high salt wash buffer.

**Table 1:**
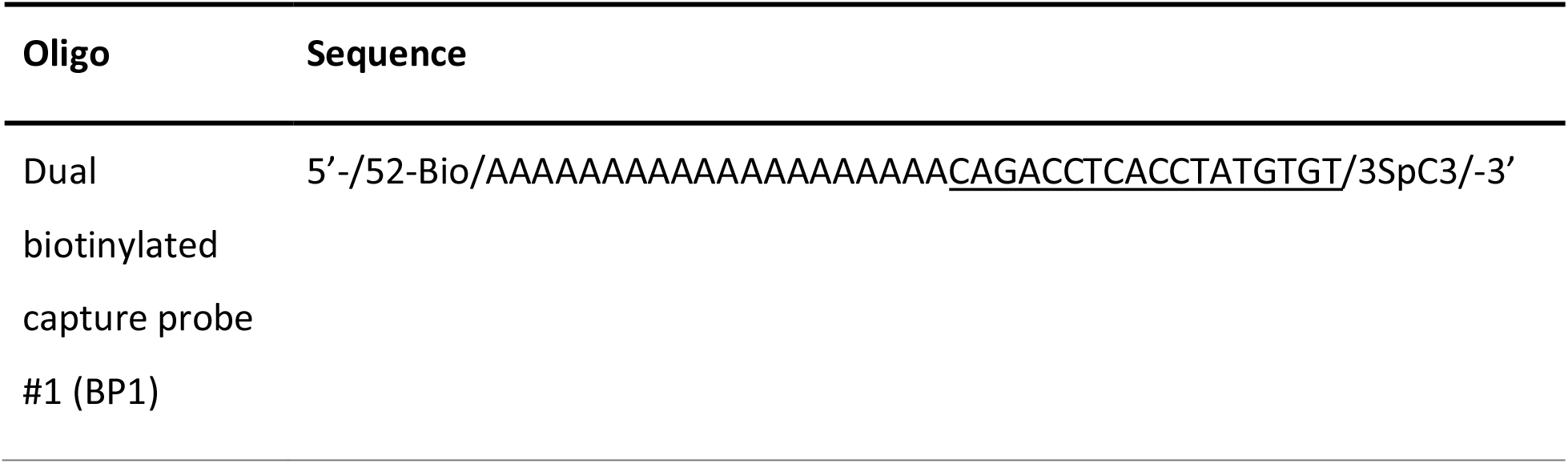
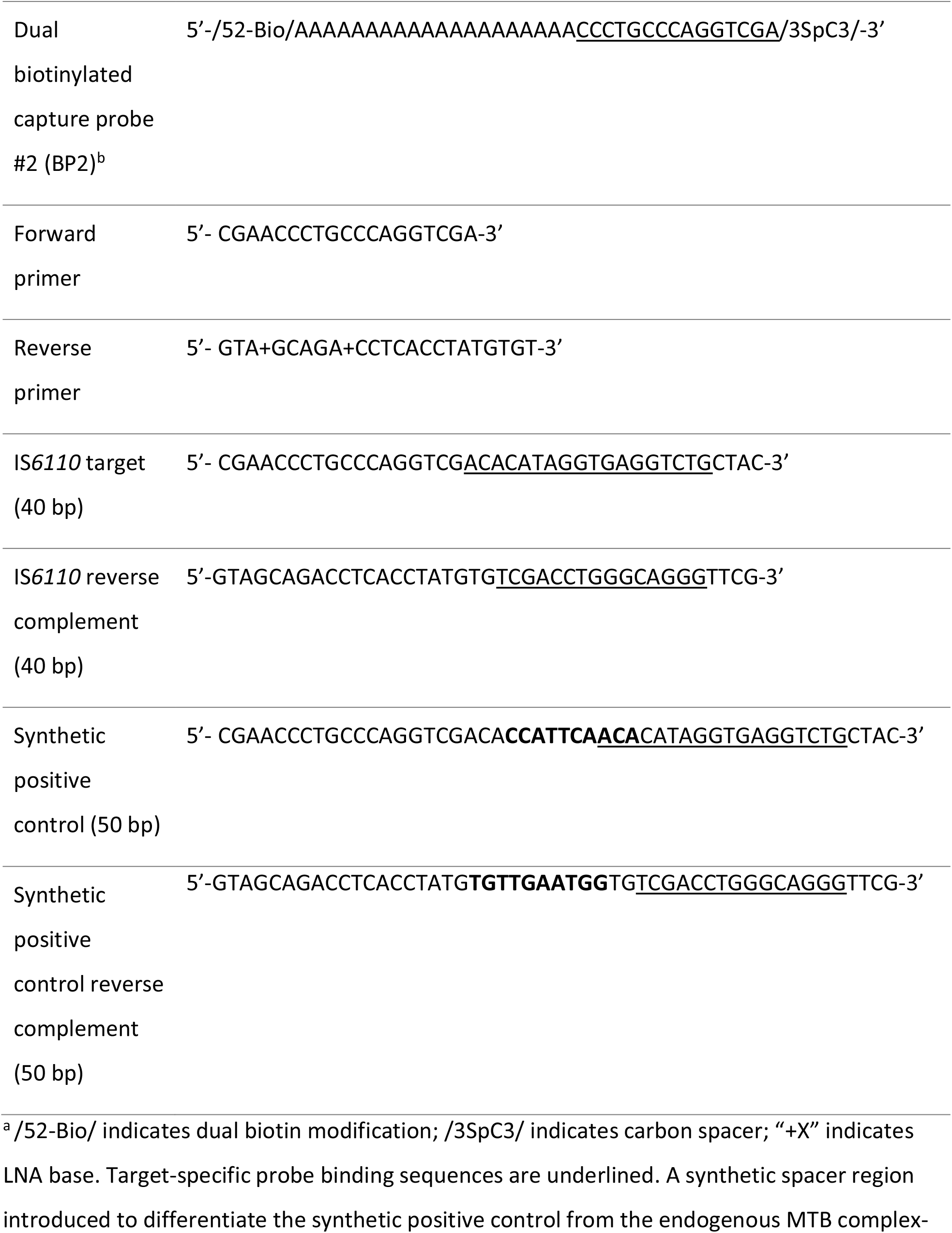
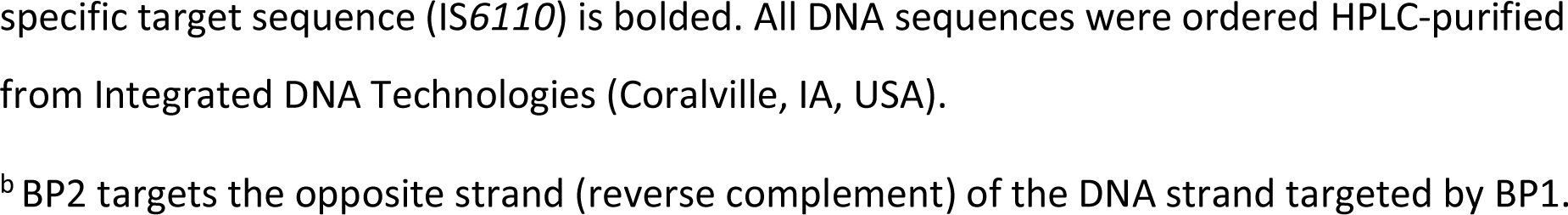
Probe, primer, and target sequences.^a^.

#### Sequence-specific capture of TB cfDNA

2.5 mL of 5 M NaCl (final concentration 1 M), 127 µL of 10% (v/v) Tween-20 (final concentration 0.1%), and 50 µL of prepared beads were added to each 10 mL urine sample and gently mixed. The samples were denatured in a heat block set to 120°C for 15 minutes, so that the urine reached a temperature of >90°C. TB cfDNA was hybridized to capture probes by rotation at room temperature for 30 minutes.

#### Washing to remove urine and non-target DNA

Samples were centrifuged for 5 minutes at 5000g to pellet beads. All but 1 mL of supernatant was removed and discarded. Beads were resuspended in the remaining supernatant and transferred to 1.5 mL DNA LoBind tubes (Eppendorf). The tubes were placed on an Invitrogen Dynamag-2 magnetic rack (Thermo Fisher) for 1 minute and the remaining supernatant was discarded. The beads were washed twice with 1 mL high salt wash buffer and once with 1 mL low salt wash buffer (15 mM NaCl, 10 mM Tris-HCl pH 8.0). For each wash, the tubes were inverted 10 – 20 times, or until no bead aggregate was left on the tube walls, briefly spun down, and placed on the magnetic rack for 1 minute before removing the wash buffer. After the final wash step, beads were spun down and any residual buffer was removed.

#### Elution of purified TB cfDNA

Purified TB cfDNA was eluted using 20 µL of freshly-prepared 20 mM NaOH. Beads were mixed with NaOH by vortexing, briefly spun down, and placed on the magnetic rack. The eluate was transferred directly to PCR wells and partially neutralized with 3.5 µL 100 mM HCl.

### Amplification of TB-specific urine cfDNA using short-target PCR

The entire output (∼24 µL) from each 10 mL urine sample was analyzed in a single PCR well. Each 50 µL reaction contained 1.25 U OneTaq Hot Start DNA Polymerase (New England Biolabs [NEB, Ipswitch, MA, USA]), 1X OneTaq GC Reaction Buffer (NEB; 80 mM Tris-SO_4_, 20 mM (NH_4_)_2_SO_4_, 2 mM MgSO_4_, 5% glycerol, 5% dimethyl sulfoxide, 0.06% IGEPAL CA-630, and 0.05% Tween-20, pH 9.2), 0.8 mM dNTPs (NEB), 0.4X EvaGreen (Biotium, Fremont, CA, USA), 200 nM forward primer (Table 1), and 200 mM reverse primer (Table 1). qPCR was carried out in a CFX96 Touch Real-Time PCR Detection System (Bio-Rad Laboratories, Hercules, CA, USA) using an initial incubation period of 94°C for 3 minutes, followed by 50 amplification cycles of 94°C for 30 seconds, 64°C for 30 seconds, and 68°C for 1 minute. Quantification cycle (C_q_) values were determined using the CFX Maestro software version 1.1 (Bio-Rad Laboratories) at a threshold of 500 RFU and recovered copies were calculated using a standard curve run with each PCR. PCR products were confirmed by post-amplification melt curve analysis from 65°C to 95°C in 0.5°C increments every 5 seconds.

### Criteria for positive samples

Criteria for positive samples were set prior to analysis, and the assay operator was blinded to the clinical status of the samples until after processing was complete and sample calls were made. Individual replicates were ruled as positive if ≥1 single-stranded DNA copy of TB cfDNA was detected and the melt temperature (T_m_) matched that of the expected native TB amplicon (76°C). Individual replicates were ruled as negative if insufficient TB cfDNA was detected (≤1 copy of single-stranded DNA) and/or the T_m_ did not match that of the expected native TB amplicon. A sample was ruled as positive if at least two of three replicates, analyzed on different days, were positive based on the above definitions. Samples with zero or one positive replicates were ruled as negative.

### PCR primer design, controls, and precautions to prevent false positives

We designed PCR primers targeting the insertion sequence IS*6110*, which is an established target for TB diagnosis present at multiple, variable copy number across ∼99% of *Mycobacterium tuberculosis* complex strains (25). Our PCR primers (Table 1) amplify a short 40 bp target within a subregion of IS*6110* that is conserved and specific to the *Mycobacterium tuberculosis* (MTB) complex (26). To avoid false positives due to nonspecific amplification, we used locked nucleic acid (LNA) bases to precisely match the T_m_ of the primers and selected an annealing temperature (T_a_) slightly above the primer T_m_ to encourage specific amplification without compromising amplification efficiency. The final primer set and optimized PCR conditions result in an average PCR efficiency of 97.1% (95% CI: 95.7 – 98.6%) and no amplification of no template controls (NTCs) or 10 ng of human genomic DNA up to at least 45 cycles. To avoid false positives due to contamination, we maintained good laboratory practices to limit contamination (*e*.*g*., separating pre- and post-PCR rooms, regular decontamination of work surfaces and pipettes, sterile filtered pipette tips, aliquoting reagents into single-use volumes). In addition, we designed the synthetic positive control (50 bp, used as a spike-in control during extraction and for PCR standard curves) to be amplifiable by the same primer pair but distinguishable from the endogenous TB target sequence (40 bp) by melt analysis (Table 1) so that any potential contamination with the positive control would not result in false positives. Every experiment included a positive control (pooled TB-negative urine spiked with 10^3^ copies of double-stranded DNA synthetic positive control template [Table 1]) and negative control (water without spiked target) that were run throughout the entire extraction process alongside clinical urine samples. PCR NTCs (n=3) were also included in each experiment.

### Statistical analysis

Sensitivity and specificity were calculated using a positive sputum Xpert MTB/RIF result and the presence of one or more TB symptoms as the reference standard. 95% confidence intervals for sensitivity and specificity were calculated using the hybrid Wilson/Brown method. Sensitivities were compared across groups using Fisher’s exact test. The detected cfDNA concentration was calculated based on the sample means of cfDNA-positive samples. Detected cfDNA concentrations were compared across groups using the Mann-Whitney test. Correlations with detected cfDNA concentration were assessed using Spearman’s correlation coefficient. Odds ratios were compared to a value of 1 using Fisher’s exact test, with 95% confidence intervals determined using the Baptista-Pike method. All statistical analysis was conducted using GraphPad Prism v8.1.2 (San Diego, CA, USA) with a significance level of 0.05.

## RESULTS

### Analytical performance of sequence-specific TB cfDNA assay

An overview of our cfDNA assay is shown in Fig. 1A-B. Recovery of the spiked-in positive control was 91.8% (95% CI: 75.9 – 107.6%) across 15 independent experiments (Fig. 1C).

**Fig. 1:**
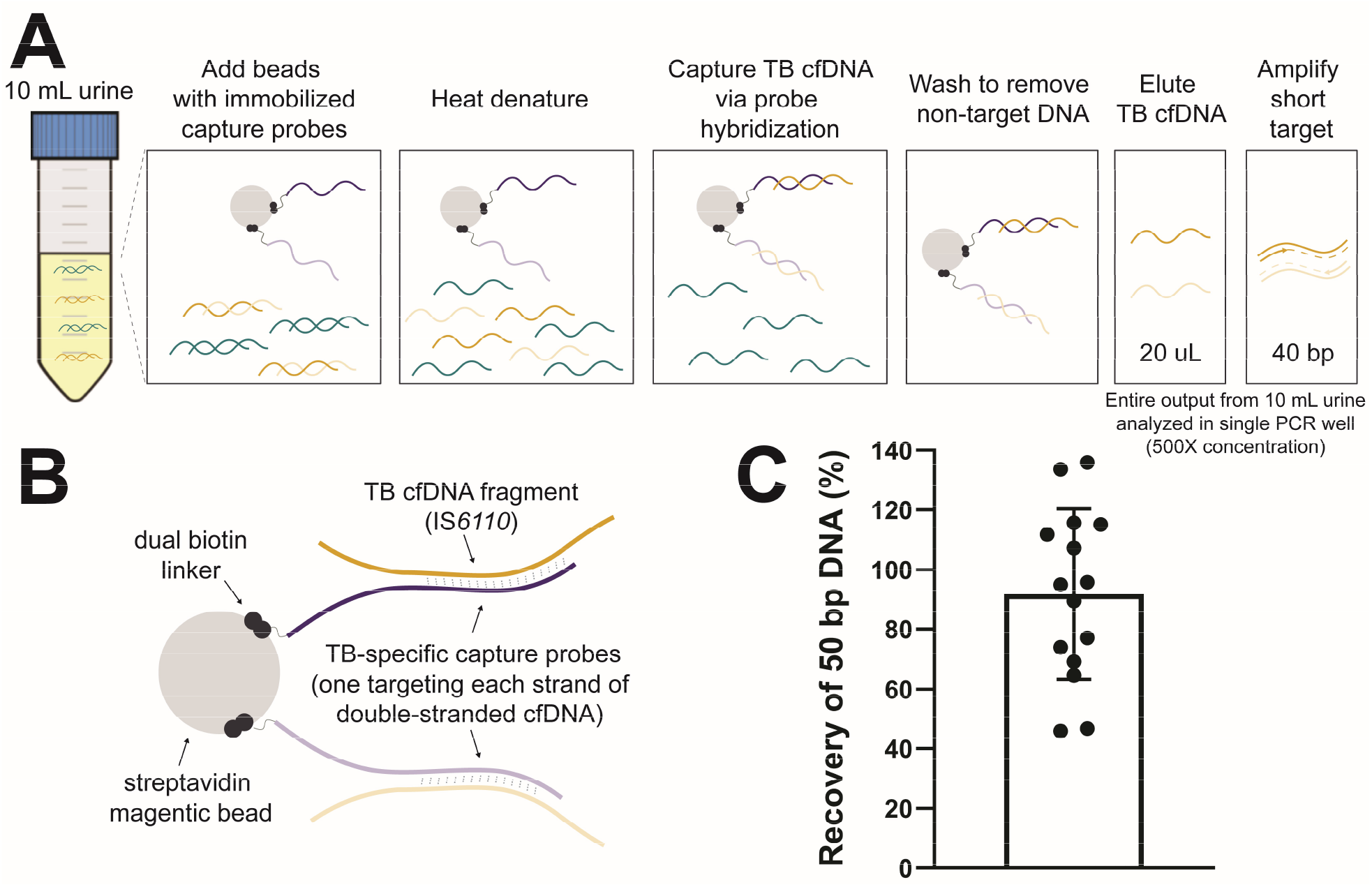
Capture and detection of short cfDNA fragments in urine using sequence-specific purification and short-target PCR. **(A)** Overview of sequence-specific purification and short-target PCR (40 bp) protocol for TB cfDNA. **(B)** Schematic illustrating details of capture probe design targeting the MTB complex-specific insertion element IS*6110*. **(C)** Near complete recovery of short TB-specific DNA spiked into urine using sequence-specific purification. A positive control (10^3^ copies of 50 bp double-stranded DNA in 10 mL pooled urine) was included throughout each experiment alongside clinical samples. The recovery of the positive control was calculated as a percentage of the input (mean ± SD, n=15). Key design features, assay optimization, and additional analytical characterization of our sequence-specific purification method for cfDNA are reported in (24).

### Study participants

We enrolled 73 participants across two sites: Edendale Hospital in South Africa (49 TB-positive, 10 TB-negative) and the University of Washington in the USA (14 healthy controls with low risk of TB exposure). Participant demographic and clinical data are summarized in Table 2.

**Table 2:**
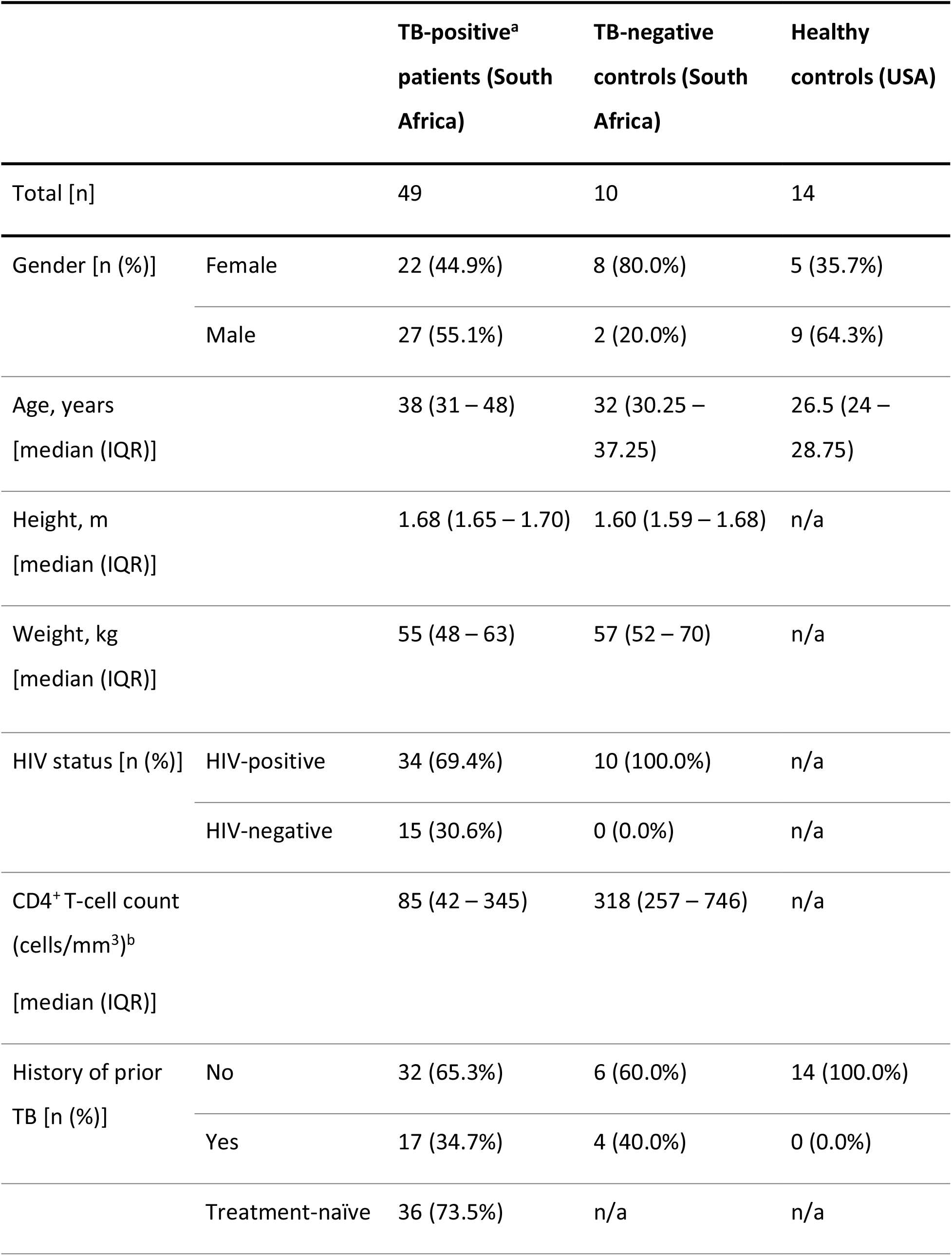
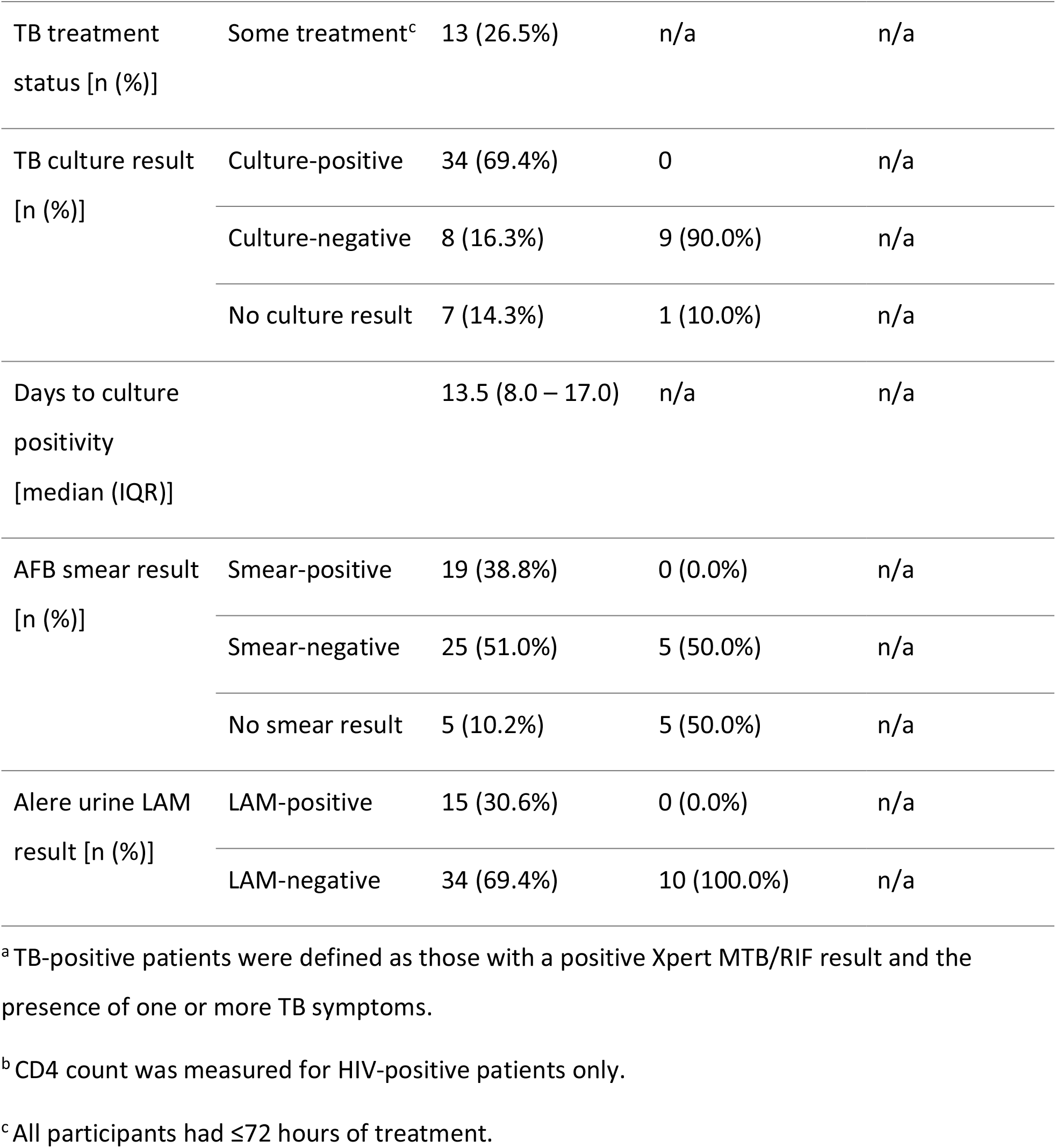
Summary of study participants.

### Sensitivity and specificity of sequence-specific TB cfDNA assay

A summary of the diagnostic accuracy of our TB urine cfDNA assay is given in Table 3. Full results for each participant, including paired clinical data and detected cfDNA concentration, are given in Supplemental Dataset S1. Using sputum Xpert MTB/RIF as the reference standard, we detected TB-specific urine cfDNA with 83.7% sensitivity (n=41/49; 95% CI: 71.0 – 91.5%) and 100% specificity (n=24/24; 95% CI: 86.2 – 100%). No TB-specific cfDNA was detected in the urine of TB-negative controls in South Africa (n=10) or healthy controls in the USA (n=14).

**Table 3:**
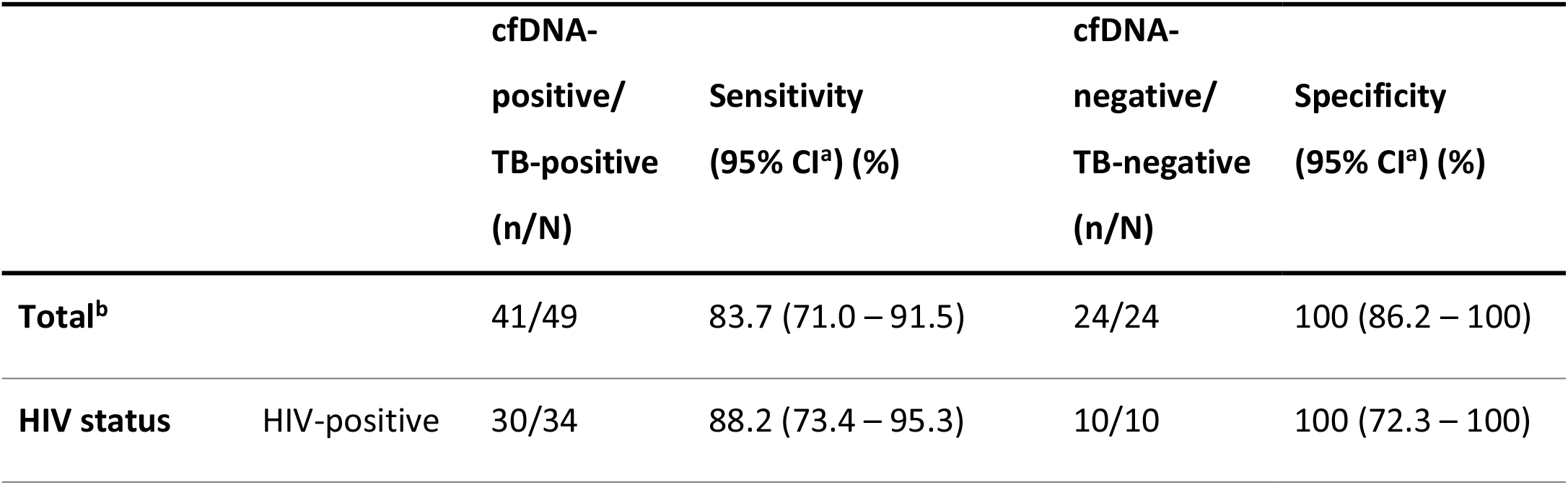
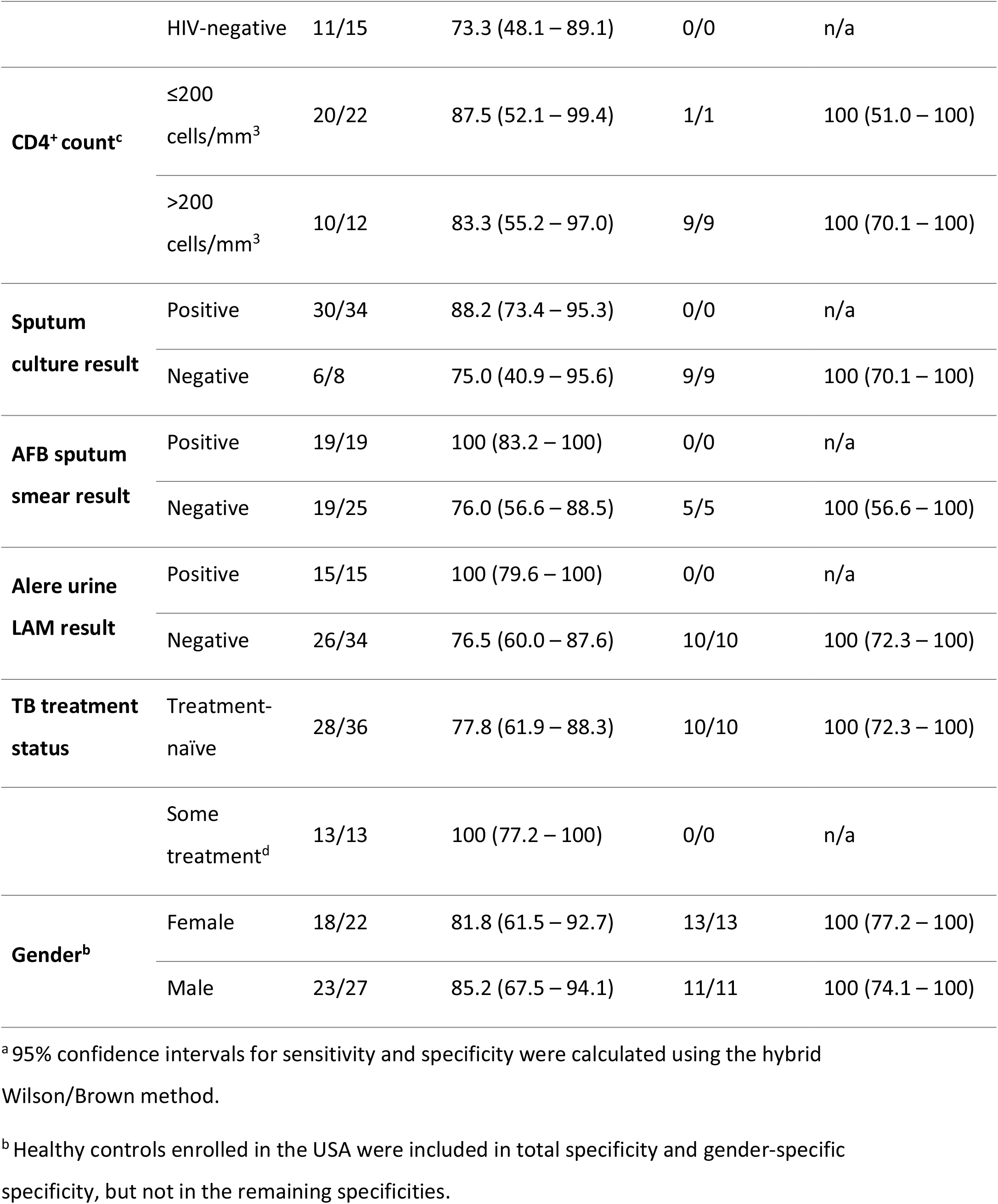
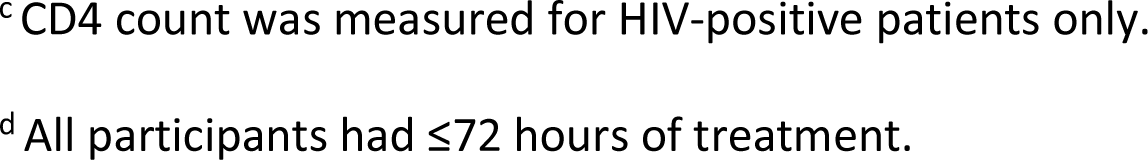
Sensitivity and specificity of TB urine cfDNA assay.

|  |  | cfDNA-positive/<br>TB-positive<br>(n/N) | Sensitivity<br>(95% CI<sup>a</sup>) (%) | cfDNA-negative/<br>TB-negative<br>(n/N) | Specificity<br>(95% CI<sup>a</sup>) (%) |
| --- | --- | --- | --- | --- | --- |
| **Total<sup>b</sup>** |  | 41/49 | 83.7 (71.0 – 91.5) | 24/24 | 100 (86.2 – 100) |
| **HIV status** | HIV-positive | 30/34 | 88.2 (73.4 – 95.3) | 10/10 | 100 (72.3 – 100) |
|  | HIV-negative | 11/15 | 73.3 (48.1 – 89.1) | 0/0 | n/a |
| **CD4<sup>+</sup> count<sup>c</sup>** | ≤200 cells/mm<sup>3</sup> | 20/22 | 87.5 (52.1 – 99.4) | 1/1 | 100 (51.0 – 100) |
|  | >200 cells/mm<sup>3</sup> | 10/12 | 83.3 (55.2 – 97.0) | 9/9 | 100 (70.1 – 100) |
| **Sputum culture result** | Positive | 30/34 | 88.2 (73.4 – 95.3) | 0/0 | n/a |
|  | Negative | 6/8 | 75.0 (40.9 – 95.6) | 9/9 | 100 (70.1 – 100) |
| **AFB sputum smear result** | Positive | 19/19 | 100 (83.2 – 100) | 0/0 | n/a |
|  | Negative | 19/25 | 76.0 (56.6 – 88.5) | 5/5 | 100 (56.6 – 100) |
| **Alere urine LAM result** | Positive | 15/15 | 100 (79.6 – 100) | 0/0 | n/a |
|  | Negative | 26/34 | 76.5 (60.0 – 87.6) | 10/10 | 100 (72.3 – 100) |
| **TB treatment status** | Treatment-naïve | 28/36 | 77.8 (61.9 – 88.3) | 10/10 | 100 (72.3 – 100) |
|  | Some treatment<sup>d</sup> | 13/13 | 100 (77.2 – 100) | 0/0 | n/a |
| **Gender<sup>b</sup>** | Female | 18/22 | 81.8 (61.5 – 92.7) | 13/13 | 100 (77.2 – 100) |
|  | Male | 23/27 | 85.2 (67.5 – 94.1) | 11/11 | 100 (74.1 – 100) |

<sup>a</sup> 95% confidence intervals for sensitivity and specificity were calculated using the hybrid Wilson/Brown method.
<sup>b</sup> Healthy controls enrolled in the USA were included in total specificity and gender-specific specificity, but not in the remaining specificities.
<sup>c</sup> CD4 count was measured for HIV-positive patients only.
<sup>d</sup> All participants had ≤72 hours of treatment.

Sensitivity was non-significantly higher in HIV-positive patients compared to HIV-negative patients (88.2% [n=30/34; 95% CI: 73.4 – 95.3%] vs. 73.3% [n=11/15; 95% CI: 48.1 – 89.1%]; p=0.23). Sensitivity was similar in HIV-positive patients with CD4 counts of ≤200 and >200 cells/mm^3^ (87.5% [n=20/22; 95% CI: 52.1 – 99.4%] vs. 83.3% [n=10/12; 95% CI: 55.2 – 97.0%]). Sensitivity was 77.8% (n=28/36; 95% CI: 61.9 – 88.3%) in treatment-naïve patients. If a positive sputum culture result was also required for the TB case definition, total sensitivity increased to 88.2% (n=30/34; 95% CI: 73.4 – 95.3%). We detected TB-specific cfDNA in the urine of all patients with positive AFB sputum smear (n=19/19) and/or Alere urine LAM (n=15/15) results. Sensitivity remained high in smear-negative (76.0% [n=19/25; 95% CI: 56.6 – 88.5%]) and LAM-negative (76.5% [n=26/34; 95% CI: 60.0 – 87.6%]) patients. The reduction in sensitivity for smear-negative patients compared to smear-positive patients was significant (p=0.029), while that for LAM-negative compared to LAM-positive patients was not (p=0.087). See Table S1 for statistical analysis of sensitivity across groups.

We evaluated factors associated with TB cfDNA positivity and found that only a positive AFB smear result was significantly associated with a positive TB cfDNA result (odds ratio >1, p=0.029). HIV status, CD4 count, treatment status, culture result, and LAM result were not significantly associated with a positive urine cfDNA result (Table S2).

### Quantification of TB-specific cfDNA concentration in urine

The detected TB-specific cfDNA concentration was variable and skewed towards low concentrations, with a median of 146 copies in 10 mL urine (Table 4). The cfDNA concentration had a moderate inverse correlation with CD4 count (−0.43 [95% CI: -0.68 to -0.10; p=0.011]) (Fig. 2A) and days to culture positivity (−0.36 [95% CI: -0.64 to -0.0060; p=0.041]) (Fig S1), but no significant correlation with days of anti-TB treatment, AFB smear score, or Alere urine LAM score (Table S3). The cfDNA concentration was higher in patients with some treatment compared to treatment-naïve patients (p=0.045) and in urine LAM-positive patients compared to urine LAM-negative patients (p=0.0045), but was not significantly different in patients grouped by HIV status, CD4 count, culture result, smear result, or gender (Fig. 2B).

**Table 4:** Detected concentrations of TB-specific urine cfDNA.

|  |  | Median (IQR)<sup>a</sup><br>[copies in 10 mL urine] | Range<sup>a</sup><br>[copies in 10 mL urine] |
| --- | --- | --- | --- |
| **Total** |  | 146 (17 – 1092) | 1.4 – 28044 |
| **TB treatment status<sup>b</sup>** | Treatment-naïve | 57 (7.6 – 557) | 1.4 – 28044 |
|  | Some treatment | 796 (104 – 2111) | 3.9 - 18543 |
| **Alere urine LAM result<sup>c</sup>** | Positive | 796 (119 – 2851) | 28 – 28044 |
|  | Negative | 39 (4.9 – 404) | 1.4 - 5021 |

<sup>a</sup> Concentration for each sample is given as copies of single-stranded DNA in 10 mL urine, which was calculated based on the mean across n=3 technical replicates.
<sup>b</sup> Detected cfDNA concentration was higher in patients with some treatment (Mann-Whitney, p=0.045).
<sup>c</sup> Detected cfDNA concentration was higher in patients with a positive urine LAM result (Mann-Whitney, p=0.0045).

**Fig. 2:**
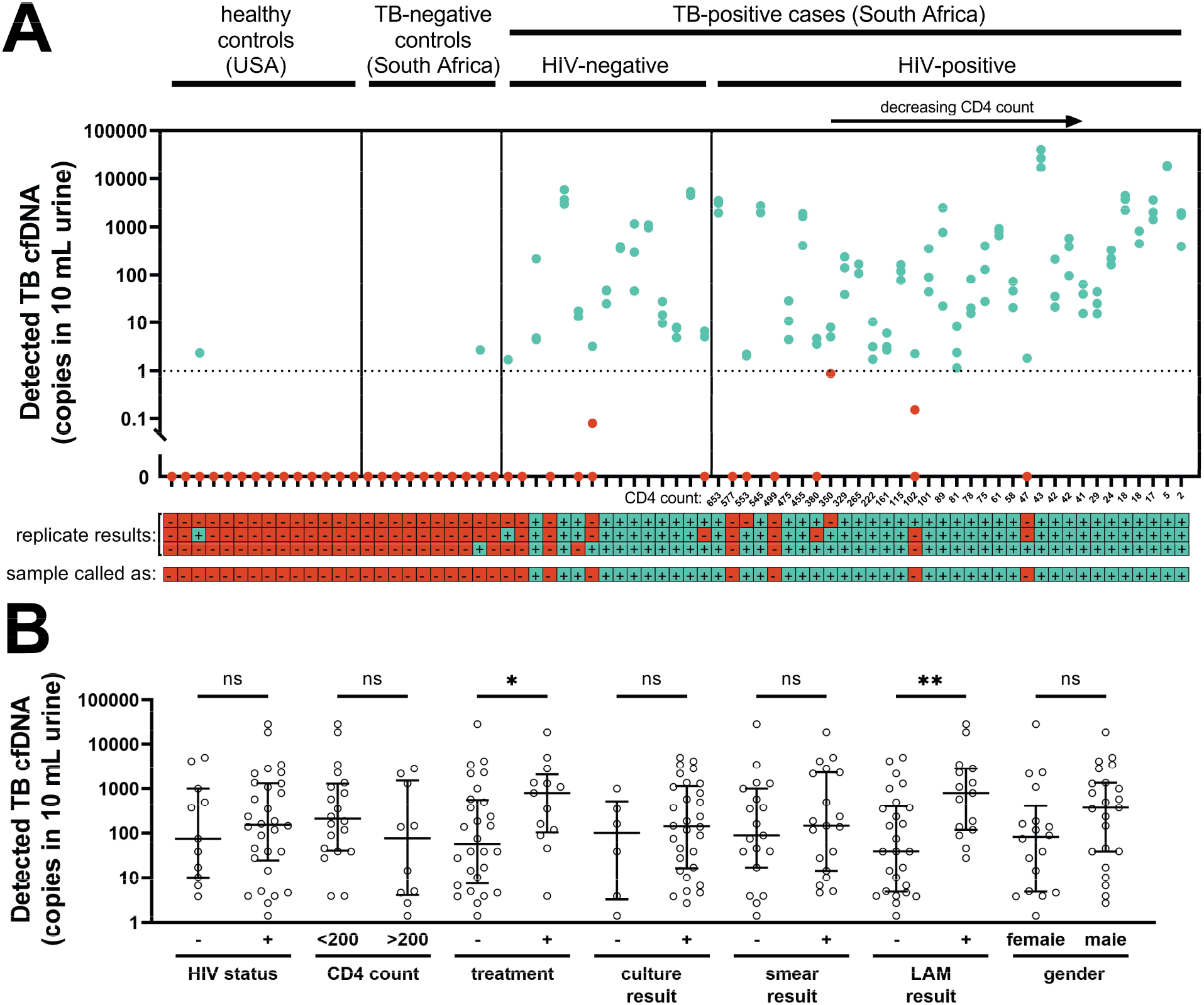
Detected concentrations of TB-specific urine cfDNA. **(A)** Concentration of TB cfDNA detected in each participant’s urine, stratified by HIV status and ranked by CD4 count. There was a moderate inverse correlation between CD4 count and detected TB cfDNA concentration (Spearman’s r = -0.43 [95% CI: -0.68 to -0.10], p = 0.011), but TB cfDNA could be detected regardless of HIV status and CD4 count. Each dot represents one of three replicates processed on different days for each sample. Note that dots representing replicates with similar detected concentrations may overlap. Replicates called as positive are shown in blue and replicates called as negative are shown in red. The dashed line indicates the 1 copy per 10 mL threshold used to define positive replicates. The legend below the plot indicates cfDNA detection status by replicate (considered positive if ≥1 copy of single-stranded DNA was detected with melt temperature matching that of the expected IS*6110* amplicon) and by sample (considered positive if ≥2 of 3 replicates were positive). **(B)** Comparison of detected TB cfDNA concentration across groups (bars indicate median and IQR of sample means of cfDNA-positive samples). The detected TB cfDNA concentration was significantly higher in patients with some treatment compared to treatment-naïve patients, and in LAM-positive patients compared to LAM-negative patients (* indicates Mann-Whitney P < 0.05; ** indicates Mann-Whitney P < 0.01), but was not affected by HIV status, CD4 count, culture result, smear result, or gender (ns indicates not significant). See Table S4 for calculated P-values for each comparison.

## DISCUSSION

### Improved detection of short cfDNA in urine using sequence-specific purification

To maximize sensitivity of TB urine cfDNA detection, it is essential to use methods designed to detect low concentrations of short fragments (6, 14, 15). Decreasing the minimum target length is expected to improve cfDNA diagnostic sensitivity (8, 27–29). For example, decreasing PCR amplicon length by a modest 10 bp (49 bp to 39 bp) led to more than 10-fold increase in detected TB-specific cfDNA (29). Critically, in addition to amplifying short targets, sample preparation methods must also extract short cfDNA from urine with high efficiency. Conventional silica-based DNA extraction methods have reduced recovery of short fragments and are thus not optimal for urine cfDNA (30).

We aimed to increase the diagnostic sensitivity of TB urine cfDNA detection by improving recovery of short cfDNA during the DNA extraction step. We developed a sequence-specific purification method that uses hybridization capture probes immobilized on magnetic beads to extract short cfDNA with high analytical sensitivity. We have previously demonstrated that sequence-specific purification improves recovery of short cfDNA fragments (25 – 150 bp) from urine compared to alternate urine cfDNA extraction methods, including a protocol used for TB urine cfDNA (31). In a recent paper detailing our sequence-specific purification method, we described its key features, provided a user-ready protocol, and thoroughly characterized its analytical performance (24). In this study, our sequence-specific approach recovered nearly all target-specific 50 bp positive control DNA (91.8% average recovery) and was in some clinical specimens able to detect down to a single copy of TB cfDNA in 10 mL urine.

### Diagnostic accuracy and comparison to previous TB urine cfDNA studies

In this study, we tested our sequence-specific TB cfDNA assay in clinical urine specimens for the first time. The sensitivity and specificity of our assay for diagnosis of active pulmonary TB were 84% and 100%, respectively, the highest reported diagnostic accuracy of a TB cfDNA test. For comparison, previous TB urine cfDNA studies are summarized in Table 5 (8–12). For a comprehensive review of previous studies, refer to (14).

**Table 5:**
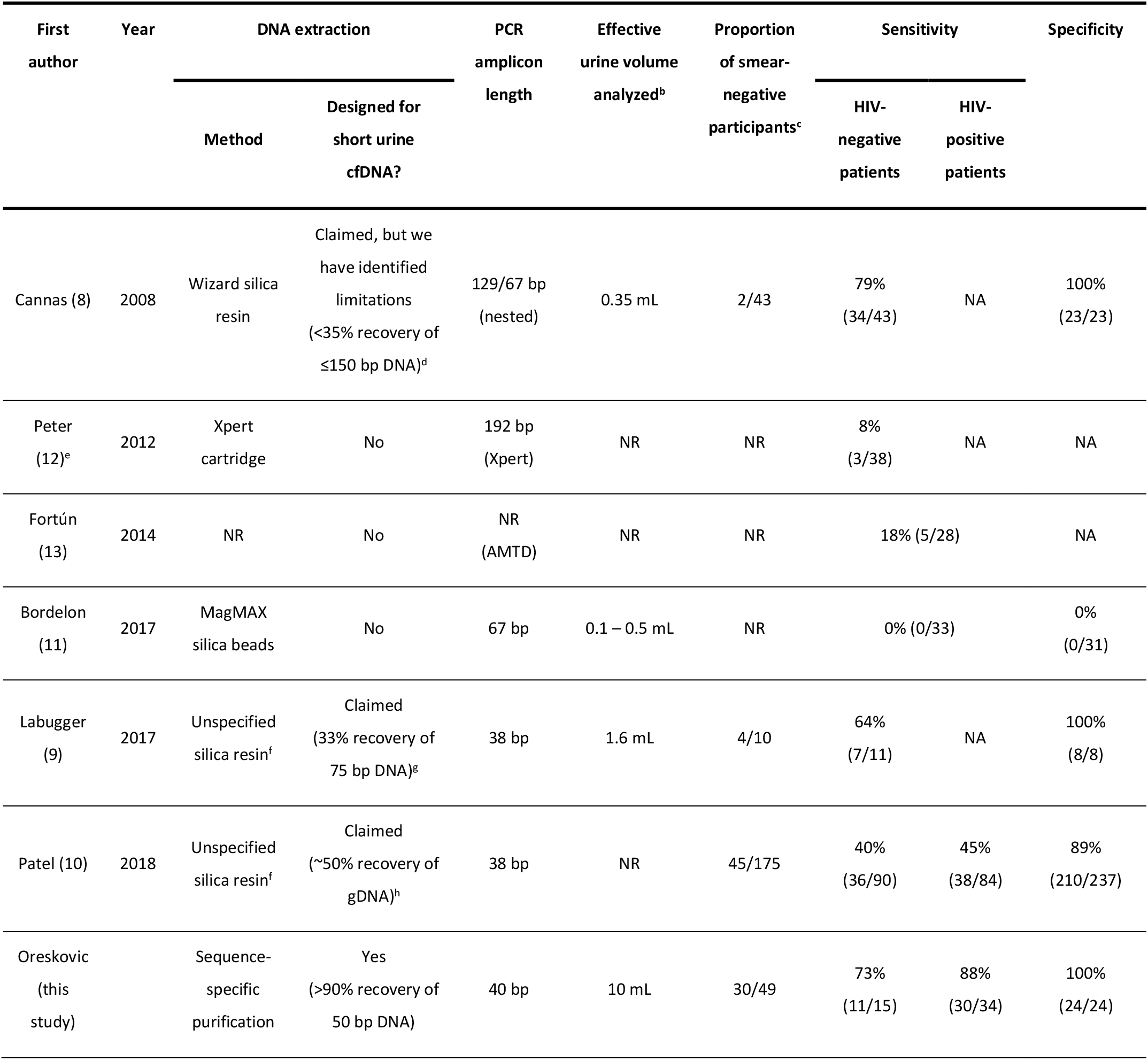
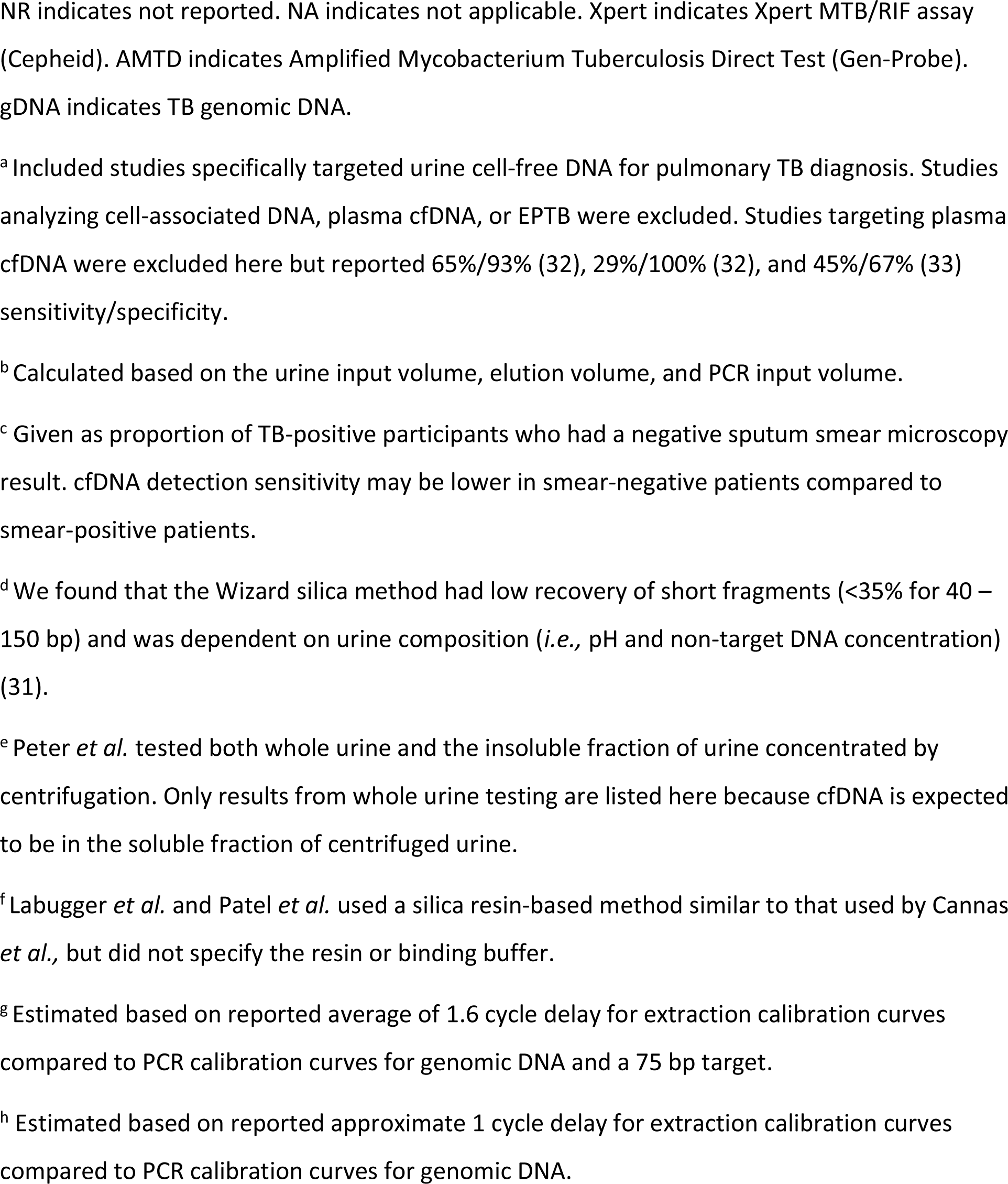
Comparison to previous studies targeting urine cfDNA for pulmonary TB diagnosis.^a^.

| First author | Year | DNA extraction |  | PCR amplicon length | Effective urine volume analyzed<sup>b</sup> | Proportion of smear-negative participants<sup>c</sup> | Sensitivity |  | Specificity |
| --- | --- | --- | --- | --- | --- | --- | --- | --- | --- |
|  |  | Method | Designed for short urine cfDNA? |  |  |  | HIV-negative patients | HIV-positive patients |  |
| Cannas (8) | 2008 | Wizard silica resin | Claimed, but we have identified limitations (<35% recovery of ≤150 bp DNA)<sup>d</sup> | 129/67 bp (nested) | 0.35 mL | 2/43 | 79% (34/43) | NA | 100% (23/23) |
| Peter (12)<sup>e</sup> | 2012 | Xpert cartridge | No | 192 bp (Xpert) | NR | NR | 8% (3/38) | NA | NA |
| Fortún (13) | 2014 | NR | No | NR (AMTD) | NR | NR | 18% (5/28) |  | NA |
| Bordelon (11) | 2017 | MagMAX silica beads | No | 67 bp | 0.1 – 0.5 mL | NR | 0% (0/33) |  | 0% (0/31) |
| Labugger (9) | 2017 | Unspecified silica resin<sup>f</sup> | Claimed (33% recovery of 75 bp DNA)<sup>g</sup> | 38 bp | 1.6 mL | 4/10 | 64% (7/11) | NA | 100% (8/8) |
| Patel (10) | 2018 | Unspecified silica resin<sup>f</sup> | Claimed (~50% recovery of gDNA)<sup>h</sup> | 38 bp | NR | 45/175 | 40% (36/90) | 45% (38/84) | 89% (210/237) |
| Oreskovic (this study) |  | Sequence-specific purification | Yes (>90% recovery of 50 bp DNA) | 40 bp | 10 mL | 30/49 | 73% (11/15) | 88% (30/34) | 100% (24/24) |

NR indicates not reported. NA indicates not applicable. Xpert indicates Xpert MTB/RIF assay (Cepheid). AMTD indicates Amplified Mycobacterium Tuberculosis Direct Test (Gen-Probe). gDNA indicates TB genomic DNA.
<sup>a</sup> Included studies specifically targeted urine cell-free DNA for pulmonary TB diagnosis. Studies analyzing cell-associated DNA, plasma cfDNA, or EPTB were excluded. Studies targeting plasma cfDNA were excluded here but reported 65%/93% (32), 29%/100% (32), and 45%/67% (33) sensitivity/specificity.
<sup>b</sup> Calculated based on the urine input volume, elution volume, and PCR input volume.
<sup>c</sup> Given as proportion of TB-positive participants who had a negative sputum smear microscopy result. cfDNA detection sensitivity may be lower in smear-negative patients compared to smear-positive patients.
<sup>d</sup> We found that the Wizard silica method had low recovery of short fragments (<35% for 40 – 150 bp) and was dependent on urine composition (*i.e.*, pH and non-target DNA concentration) (31).
<sup>e</sup> Peter *et al.* tested both whole urine and the insoluble fraction of urine concentrated by centrifugation. Only results from whole urine testing are listed here because cfDNA is expected to be in the soluble fraction of centrifuged urine.
<sup>f</sup> Labugger *et al.* and Patel *et al.* used a silica resin-based method similar to that used by Cannas *et al.*, but did not specify the resin or binding buffer.
<sup>g</sup> Estimated based on reported average of 1.6 cycle delay for extraction calibration curves compared to PCR calibration curves for genomic DNA and a 75 bp target.
<sup>h</sup> Estimated based on reported approximate 1 cycle delay for extraction calibration curves compared to PCR calibration curves for genomic DNA.

Prior studies showed potential for detection of TB-specific urine cfDNA in HIV-positive and HIV-negative patients, but had variable sensitivity due to sample preparation and/or amplification methods sub-optimal for short targets (6, 14, 15). Cannas *et al*. used an in-house silica resin method (based on Promega Wizard DNA Purification Resin) reportedly designed to improve binding of short cfDNA, but did not report its analytical performance (*e*.*g*., percent recovery) (8). While the Wizard method improves upon conventional silica-based methods, our past testing revealed limited recovery (<35%) of ≤150 bp fragments (31). It was also highly dependent on urine composition and is likely to fail in samples with high pH and/or low non-target DNA concentrations (31). Labugger *et al*. and Patel *et al*. used a similar silica resin-based method, but did not specify the resin or binding conditions (9, 10). It is possible that their method suffers from similar limitations as the Wizard method used by Cannas *et al*., although we could not experimentally verify this. They reported recoveries of approximately 30 – 50% and limits of detection of 3 copies/mL (75 bp target) (9) and 1.25 copies/mL (TB gDNA containing up to 42 target repeats) (10). Our sequence-specific purification method increases percent recovery by 2-fold or more and improves the limit of detection to ≤0.5 copies/mL of 50 bp cfDNA (positivity cutoff threshold of 0.1 copies/mL used here). Our method allows the full volume eluted from a 10 mL urine extraction to be amplified in a single PCR well, maximizing sensitivity for detection of low-concentration samples.

Past studies also enrolled few smear-negative participants, and thus have not tested the ability to detect urine cfDNA in individuals with paucibacillary TB who stand to benefit most from a non-sputum-based test. In particular, the most promising study by Cannas *et al*. included only 5% (2/43) smear-negative participants. Our results indicate that cfDNA detection sensitivity is higher in smear-positive compared to smear-negative individuals, so enrollment biased towards smear-positive participants may lead to an overestimation of assay sensitivity. In contrast, we demonstrated higher sensitivity while including 61% (30/49) smear-negative participants.

Although not directly tested here, an added benefit of our sequence-specific approach is that it may help ensure specificity by removing non-target DNA. Sequence-specific purification has been used to improve sensitivity of TB diagnosis from sputum by removing high concentrations of non-target DNA that can lead to downstream amplification inhibition (34), but has not been applied in urine where the primary advantage of non-target DNA removal would be to reduce the likelihood of downstream nonspecific amplification. Confirming specific amplification of short targets without the footprint for a fluorescent detection probe can be difficult, but the added layer of specificity offered by sequence-specific purification may aid in overcoming this challenge.

### Comparison to existing rapid TB tests

Our urine cfDNA assay has the potential to diagnose TB in individuals who may be missed by other rapid tests (*e*.*g*., smear microscopy, urine LAM). We detected TB-specific cfDNA in the urine of all smear-positive and LAM-positive patients. Importantly, sensitivity remained high in both smear-negative (76.0%) and LAM-negative (76.5%) patients. The target product profile for a rapid biomarker-based non-sputum-based TB test outlines the optimal requirements for pulmonary TB in adults as ≥98% for smear-positive TB, ≥68% for smear-negative TB, and ≥80% sensitivity for HIV-associated TB (5). Our results suggest that, by employing an optimized extraction method with demonstrated high efficiency for short fragments, urine cfDNA-based assays have the potential to achieve sufficient sensitivity to meet these criteria and improve diagnostic accuracy compared to smear microscopy.

Unlike urine LAM tests, which have insufficient sensitivity, particularly in HIV-negative individuals (35), cfDNA tests have the potential to diagnose TB regardless of HIV status and CD4 count. We observed slightly higher sensitivity in HIV-positive (88.2%) compared to HIV-negative (73.3%) patients, but the difference was non-significant and the number of HIV-negative patients was small. Despite a moderate inverse correlation between CD4 count and detected TB-specific cfDNA concentration, there was no difference in sensitivity for HIV-positive patients with CD4 counts of ≤200 compared to >200 cells/mm^3^. In contrast, the commercially-available Alere Determine TB LAM test has 42% pooled sensitivity in HIV-positive individuals, with sensitivity inversely proportional to CD4 cell count (36). The WHO only recommends its use in people living with HIV but not as a general screening test for TB (37). Ongoing efforts aim to improve urine LAM detection sensitivity. The Fujifilm SILVAMP TB LAM test improves sensitivity relative to Alere LAM in both HIV-positive (70% vs. 42%) and HIV-negative (53% vs. 11%) individuals (38, 39). As antigen-based tests, however, LAM assays may not be able to achieve the sensitivity afforded by nucleic acid amplification tests.

### Contributions to evidence for urine cfDNA as a biomarker for TB

To date, usefulness of urine cfDNA as a biomarker for TB has been limited, in part due to inconsistent methods and results in previous TB urine cfDNA studies. Our study contributes to the evidence for urine cfDNA as a TB biomarker in two ways: 1) demonstration of the feasibility of high sensitivity and specificity with optimal pre-analytical methods and 2) development of a reliable, quantifiable method to further study TB urine cfDNA. Previous TB urine cfDNA studies have focused on measuring diagnostic accuracy, and have mostly neglected to report TB urine cfDNA concentrations. Labugger *et al*. measured concentrations for a limited number of treatment-naïve individuals (n=7), which ranged from 1 – 41 copies/mL (median 6.5 copies/mL) (9). Here, we quantified TB-specific cfDNA to better estimate the clinical range (<1 – 2804 copies/mL; median 14.6 copies/mL), and have conducted analyses to determine which variables correlate with cfDNA concentration. We found that cfDNA concentration was higher in LAM-positive patients compared to LAM-negative patients, but LAM result did not affect cfDNA sensitivity. We detected cfDNA in all patients who had recently initiated TB treatment, with a higher concentration compared to treatment-naïve patients, supporting the possibility of using cfDNA for treatment monitoring as suggested previously (9). Upon initiation of a successful treatment regimen, cfDNA concentration may temporarily increase, followed by a slow decline as the infection is cleared (9). Our study did not show a correlation between days of treatment and cfDNA concentration, but only included participants with ≤3 days of treatment and did not monitor individual participants over time.

Although cfDNA was detectable regardless of HIV status or CD4 count, the detected concentration had a moderate inverse correlation with CD4 count. We also found that cfDNA concentration had a moderate inverse correlation with days to culture positivity, suggesting that levels of excreted cfDNA may be related to bacterial burden. We observed no correlation with AFB sputum smear score or Alere urine LAM score, but the sample sizes for these analyses were low. We anticipate that our assay can be used to continue to study TB urine cfDNA trends across sub-groups and answer important unresolved questions regarding optimal sample collection techniques, processing methods, and storage conditions that have been the focus of recent work (40–42). As a caveat, the cfDNA concentrations measured by our assay may be confounded by the variable copy number of IS*6110* (0–25 copies) (25) and by differences in participants’ hydration statuses. In the future, strain typing and normalizing cfDNA concentration to urine creatinine may help better elucidate trends in cfDNA concentration.

### Study limitations and future work

TB symptoms were required in addition to a positive Xpert result for the TB case definition to reduce the risk of a false-positive Xpert result, but it is possible that some could still have occurred. Using the strictest TB-positive criteria, requiring both positive Xpert and positive culture, would result in a non-significant increase in cfDNA sensitivity (88%). Several TB-positive, cfDNA-negative samples (false negatives) narrowly missed the cfDNA positivity cutoff, suggesting the opportunity for future improvement in clinical sensitivity. We are currently pursuing two approaches to further improve our assay’s ability to detect low cfDNA concentrations: a reduction in PCR target length (to 25 bp using an ultrashort PCR design described in (31)) and multiplexing to target multiple genomic regions. Multiplexing will have the additional benefit of improving inclusivity by enabling detection of TB strains lacking IS*6110*.

This study serves as a valuable demonstration of the feasibility of our sequence-specific approach, but the sample size and scope were limited. Future studies will seek to further validate our assay with larger sample sizes (including more HIV-negative participants and negative controls) and expanded populations, specifically those underserved by current rapid sputum-based tests (including children and patients with EPTB). TB-specific cfDNA has been detected in the urine (13) and plasma (43–45) of individuals with EPTB, including a case study of a child with tubercular otitis media (46), but there have not yet been any prospective studies in children. Our assay also currently requires a trained user and laboratory equipment. In the future, we aim to simplify our lab-based test into a format more suitable for use in resource-limited settings.

## Conclusion

Sequence-specific purification improves recovery of short urine cfDNA and increases the sensitivity of TB diagnosis from urine cfDNA. Our assay has the highest reported accuracy of any TB urine cfDNA test to date and has the potential to enable urine-based TB diagnosis across sputum-scarce and paucibacillary populations. This study will lay the foundation for expanded clinical studies and future development of a rapid test. In addition, our work serves as a valuable contribution to the clinical evidence for urine cfDNA as a biomarker for TB. The ability to diagnose TB across key underserved populations (*e*.*g*., children, people living with HIV, individuals with EPTB) using urine samples would address an urgent need that was identified as one of the highest priority gaps in TB diagnostics (5). A sensitive urine-based test built upon the sequence-specific purification method described here could significantly contribute to improving sample availability, expanding access to rapid TB diagnosis, and controlling the TB epidemic.

## Data Availability

All relevant data are within the paper and its Supporting Information files.

## ACKNOWLEDGEMENTS

We are grateful to study participants for their contributions to this research. We thank the KwaZulu-Natal Department of Health and staff of Edendale Hospital for their partnership and Norman D. Brault and James Lai for their contributions to the early development of the sequence-specific purification method. Research reported in this publication was funded by the Bill and Melinda Gates Foundation under award number OPP1152864 and the National Institute of Allergy and Infectious Diseases of the National Institutes of Health under award number R21AI125975. The content is solely the responsibility of the authors and does not necessarily represent the official views of the National Institutes of Health. This research was funded in part by a 2018 CFAR developmental grant from the University of Washington/Fred Hutch Center for AIDS Research, an NIH-funded program under award number AI027757 which is supported by the following NIH Institutes and Centers: NIAID, NCI, NIMH, NIDA, NICHD, NHLBI, NIA, NIGMS, NIDDK. A.O. was supported by funding from the National Science Foundation Graduate Research Fellowship Program. The funders had no role in study design, data collection and interpretation, or the decision to submit the work for publication.

