## Supplemental Information for "Diagnosing pulmonary tuberculosis using sequence-specific purification of urine cell-free DNA"

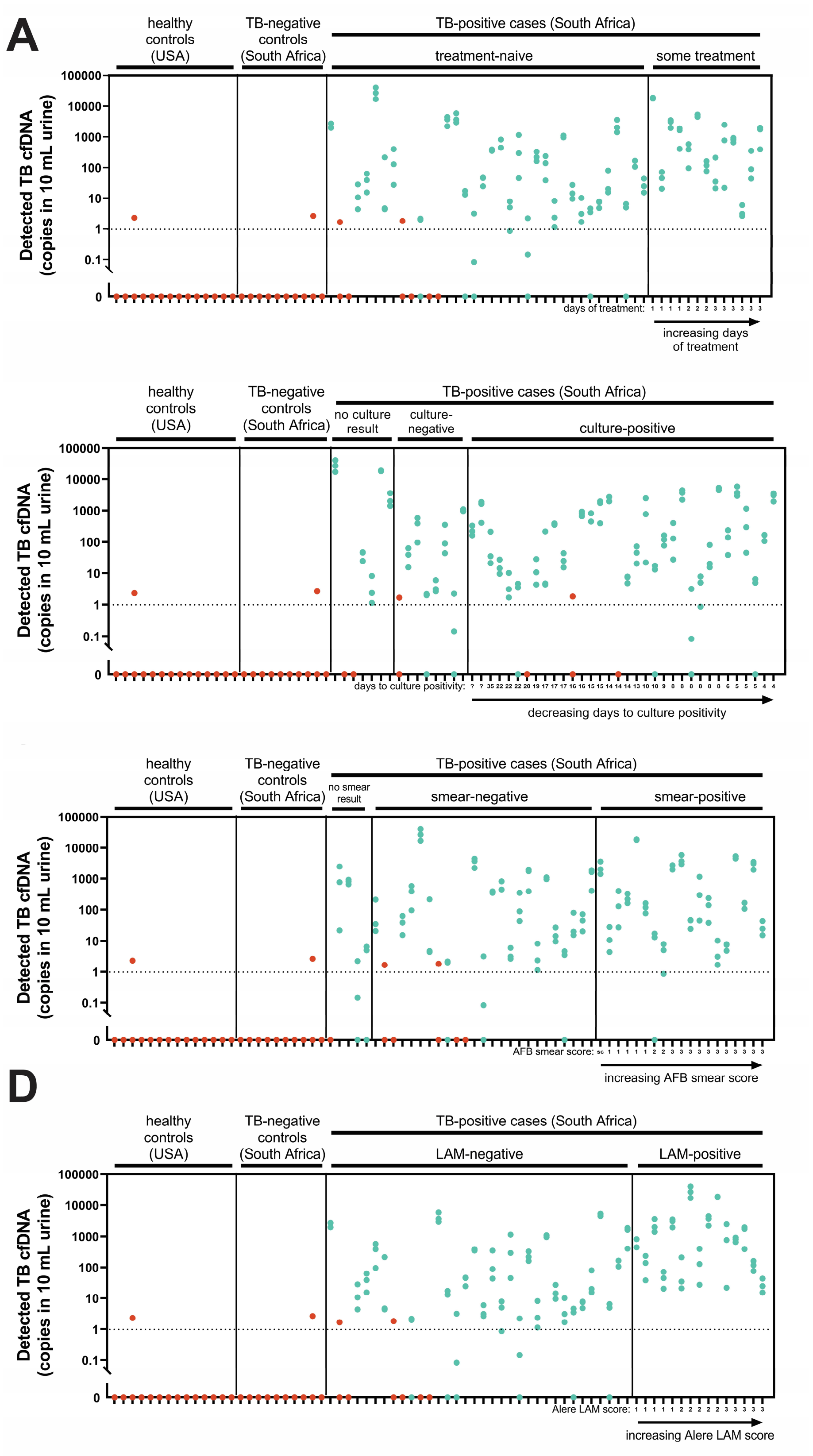


**Supplemental Fig. S1: Detected TB-specific urine cfDNA concentrations.** For all plots, each dot represents one of three replicates per sample, processed on different days. Blue dots represent positive replicates with ≥1 copy detected and melting temperature matching the expected IS*6110* amplicon. Red dots represent negative replicates. At least two out of three replicates were required to be positive for a sample to be called as positive. **(A)** Stratified by treatment status and ranked by days of treatment (no correlation). **(B)** Stratified by culture result and ranked by days to culture positivity (Spearman r = -0.36 [95% CI: -0.64 to -0.0060], p = 0.041). **(C)** Stratified by smear result and ranked by smear score (no correlation). **(D)** Stratified by Alere urine LAM result and ranked by Alere urine LAM score (no correlation).

**Supplemental Table S1: Comparison of sensitivities across groups.**

| Variable | Groups for comparison of sensitivity | P-value[a] |
|---|---|---|
| **HIV status** | positive vs negative | 0.23 |
| **CD4 count[b]** | ≤200 vs >200 cells/mm$^3$ | >0.99 |
| **TB treatment status** | treatment-naïve vs some treatment | 0.090 |
| **Sputum culture result** | positive vs negative | 0.32 |
| **AFB sputum smear result** | positive vs negative | 0.029* |
| **Alere urine LAM result** | positive vs negative | 0.087 |
| **Gender** | female vs male | >0.99 |

* Indicates P < 0.05

[a] P-values comparing sensitivity across groups were calculated using Fisher's exact test for relative risk ratios.

[b] CD4 count was measured for HIV-patients only.

**Supplemental Table S2: Diagnostic odds ratios indicating associations with a positive urine cfDNA result.**

| Variable | Comparison | Odds ratio (95% CI)[a] | P-value[b] |
|---|---|---|---|
| **HIV status** | positive vs negative | 2.7 (0.68 – 10.5) | 0.23 |
| **CD4 count[c]** | ≤200 vs >200 cells/mm$^3$ | 2.0 (0.28 – 13.9) | 0.60 |
| **TB treatment status** | treatment-naïve vs some treatment | infinity (0.79 – infinity) | 0.090 |
| **Sputum culture result** | positive vs negative | 2.5 (0.39 – 14.7) | 0.32 |
| **AFB sputum smear result** | positive vs negative | infinity (1.6 – infinity) | 0.029* |
| **Alere urine LAM result** | positive vs negative | infinity (1.0 – infinity) | 0.087 |

* Indicates P < 0.05

[a] Diagnostic odds ratio of >1 indicates variables associated with higher likelihood of a positive cfDNA result. 95% confidence intervals calculated using the Baptista-Pike method.

[b] Diagnostic odds ratios were compared to a value of 1 using Fisher's exact test.

[c] CD4 count was measured for HIV-patients only.

**Supplemental Table S3: Correlations with detected TB-specific cfDNA concentration.**

| Variable | Spearman's rank correlation coefficient (95% CI) | P-value |
|---|---|---|
| **CD4 count[a]** | -0.43 (-0.68 to -0.10) | 0.011* |
| **Days of TB treatment[b]** | -0.36 (-0.77 to 0.26) | 0.23 |
| **Days to sputum culture positivity** | -0.36 (-0.64 to -0.0060) | 0.041* |
| **AFB sputum smear score[c]** | -0.37 (-0.77 to 0.24) | 0.21 |
| **Alere urine LAM score[d]** | -0.094 (-0.59 to 0.45) | 0.76 |

* Indicates P < 0.05

[a] CD4 count was measured for HIV-patients only.

[b] Correlation calculated only for patients with some treatment (1 – 3 days). Treatment-naïve patients were excluded.

[c] Correlation calculated only for smear-positive patients (AFB score ≥1). Smear-negative patients were excluded.

[d] Correlation calculated only for LAM-positive patients (Alere LAM score ≥1). LAM-negative patients were excluded.

**Supplemental Table S4: Comparison of detected TB-specific cfDNA concentration across groups.**

| Variable | Groups for comparison of detected cfDNA concentration | P-value[a] |
|---|---|---|
| **HIV status** | positive vs negative | 0.8007 |
| **CD4 count[b]** | ≤200 vs >200 cells/mm$^3$ | 0.1946 |
| **TB treatment status** | treatment-naïve vs some treatment | 0.0447* |
| **Sputum culture result** | positive vs negative | 0.3709 |
| **AFB sputum smear result** | positive vs negative | 0.7013 |
| **Alere urine LAM result** | positive vs negative | 0.0045** |
| **Gender** | female vs male | 0.1081 |

* Indicates P < 0.05; ** Indicates P < 0.01

[a] P-values calculated using Mann-Whitney test comparing sample means of cfDNA-positive samples.

[b] CD4 count was measured for HIV-patients only.
